# Acute effects of the 1-minute sit-to-stand test (STST) on immune-metabolic stress indices

**DOI:** 10.64898/2026.03.05.26347678

**Authors:** Wiebke Röhr, Rebecca Simon, Sebastian Kirschke, Isabell von Loga, David Putrino, Wilhelm Bloch, Philipp A. Reuken, Diana Dudziak, Anna P. Kipp, Andreas Stallmach, Christian Puta

**Author notes:** Corresponding author Contact information: Department of Sports Medicine and Health Promotion Friedrich Schiller University Jena Wöllnitzer Straße 42 07749 Jena, Germany.

## Abstract

Characterizing the non-pathological, exercise-induced immune-metabolic stress regulation is important for understanding physiological adaptations and distinguishing them from maladaptive responses. Because such responses are mostly studied with extensive, resource-intensive protocols, simple, standardized tests eliciting measurable immune-metabolic responses are valuable in clinical and non-clinical settings. Established field tests assess functional exercise capacity rather than providing a standardized stimulus for investigating exercise-induced immune regulation. This study examined the effects of the 1-minute sit-to-stand test (STST) on immune-metabolic stress indices and whether it elicits sufficient anaerobic stress to serve as a tool for investigating exercise-induced immunological stress responses. Capillary blood from 28 healthy adults was collected before, immediately after, and during 45 minutes after exercise to determine lactate, glucose, and blood cell counts; hemoconcentration correction was applied. Lactate increased significantly immediately after the STST, exceeding the anaerobic threshold (>4 mmol/L). Leukocyte, lymphocyte, and granulocyte counts rose significantly post-exercise; leukocytes returned to baseline by 30 minutes, whereas granulocytes remained elevated. GLR and SII decreased significantly immediately post-exercise and increased during recovery, together with SIRI. Lactate changes correlated significantly with leukocyte, lymphocyte and granulocyte changes. Therefore, the STST represents a practical, standardized field-based model for immune-metabolic assessment with translational potential for clinical and non-clinical settings.

## 1 Introduction

Exercise is increasingly recognized as a potent modulator of immune function, influencing immune cell mobilization, phenotype, and function through coordinated metabolic, endocrine, and inflammatory pathways [1]. Acute exercise induces a local and systemic response characterized by activation of the immune and inflammatory systems immediately after the activity [2,3], though the magnitude of this response is often dependent on parameters related to the physical activity (type, duration, intensity and frequency) [4]. During and after exercise, proinflammatory cytokines, especially interleukin-6 (IL-6) and tumor necrosis factor-α (TNF-α), are released first, and anti-inflammatory cytokines such as IL-4 or IL-10 are released shortly thereafter [2,4,5]. Mediated by the cytokine release, acute phase proteins (APP), such as C-reactive protein (CRP) will also subsequently and temporarily rise (8 to 12 hours later) [5]. However, long-term regular physical activity and good aerobic fitness are associated with a reduction in inflammatory markers such as CRP [6,7].

For aerobic and anaerobic physical exertion of less than 2 hours, the body’s immunological reaction can be divided into 2 phases: immediate regulation [8] and delayed regulation [9]. Immediate regulation occurs during and immediately after physical exertion and primarily represents a stress response of the sympathetic nervous system [10,11]. This leads to the production and release of catecholamines, namely adrenaline and noradrenaline, via the adrenal medulla (sympathetic nervous system axis), whereby stress-relevant functions are promoted within seconds through the activation of α- and β-adrenergic receptors [10,11]. Subsequently, heartbeat and breathing rate increase, blood flow to large muscle groups is increased and energy reserves are mobilized [11]. Delayed regulation starts 30 minutes to one hour after exercise and represents the slow stress response, which is characterized by the release of the glucocorticoid cortisol from the adrenal cortex (hypothalamic-pituitary adrenal axis) [8,10]. Catecholamine and glucocorticoid release contribute to the distribution of immune cells between blood and peripheral pools [3,12]. The release of catecholamines during exercise results in a rapid increase in circulating leukocyte count [3,10]. About half of this increase is due to the increasing number of granulocytes in the blood [9]. However, the proportion of granulocytes decreases over time due to lower catecholamine sensitivity compared to lymphocytes and monocytes [9]. There is an increase in the number and activity of lymphocytes (lymphocytosis) [10], whereby increased shear forces and blood pressure also lead to their mobilization as a result of the activity [13]. Glucocorticoids, such as cortisol, influence key mechanisms underlying leukocyte diapedesis by regulating various facets of endothelial physiology [14]. A delayed stress-induced regulation, characterized by reduced lymphocyte counts and neutrophilia, can be observed after exercise [3,10]. Lymphocyte and monocyte counts return to or even fall below baseline levels 30 minutes to 2 hours after the end of exercise [3,8].

The predominant metabolic pathway for energy production during physical exertion depends on the intensity and duration of the exercise. At the onset of intense short-term exercise, both anaerobic and aerobic metabolic pathways are activated [15]. However, due to their high energy turnover rates, anaerobic metabolic pathways enable faster ATP provision compared to aerobic pathways, but the ATP yield is considerably lower [15,16]. During anaerobic activities lasting a few seconds to several minutes, ATP is primarily generated through the breakdown of glucose and glycogen via anaerobic glycolysis, resulting in lactate production [15]. When high-intensity exercise is sustained, lactate accumulates in the working muscles and is subsequently released into the bloodstream [17,18]. Consequently, blood lactate concentration serves as a key indicator for determining the anaerobic threshold. This threshold marks the increasing shift toward anaerobic glycolysis with lactate accumulation, due to a relative insufficiency of oxygen delivery to meet muscular energy demands [19].

However, especially the exercise-related alterations concerning the immune system have been documented primarily in studies using extensive, high-infrastructure and resource-intensive exercise protocols (e.g. graded ergometer tests, maximal exercise tests, venous blood sampling) [9,20–22]. For clinical and non-clinical settings, there is a critical need to better characterize the immune-metabolic stress regulations that occur in response to acute short-term exercise bouts. The metabolic milieu generated during high-intensity exercise exerts direct modulatory effects on immune cell function [23,24]. Skeletal muscle contraction during anaerobic metabolism results in the accumulation of lactate and hydrogen ions, producing a transient metabolic acidosis [16]. Blood lactate concentrations exceeding 4 mmol/L, the conventional threshold demarcating the transition to predominantly anaerobic metabolism [25], are therefore associated with altered immune cell function through multiple mechanisms. The application of simple, standardized exercise protocols capable of eliciting measurable immune-metabolic responses in field and clinical settings addresses a critical methodological gap. Such protocols would enable longitudinal monitoring of immune function in preventive and rehabilitative contexts, facilitate early detection of maladaptive responses, and provide objective outcome measures for interventional studies targeting immune-metabolic health.

Established simple field tests used for the assessment of exercise tolerance and physical performance in different pathological and non-pathological settings are timed walk tests (e.g., 6-minute walk test) and step tests (e.g., chester step test). These tests represent protocols that involve lower costs, fewer equipment requirements and easier administration than the commonly used tests with cycle ergometer or treadmill. The 6-minute walk test requires a 30 m-hallway and is generally recommended to be preceded by a familiarization trial, while the chester step test is physically demanding and cannot be completed by some individuals. Therefore, the 1-minute sit-to-stand test (STST), another simple field test that is characterized by ease of administration, availability of equipment and low costs, represents an alternative to those tests [26]. Compared to the other two tests, the STST has the advantages that even individuals with limited physical fitness can complete it and it requires only small spaces and less time which highlights the potential clinical applications of this test. Unlike the other tests, the STST can therefore be used in a greater variety of settings and locations in- and outside the hospital, whereby potential applications in primary care and tele-rehabilitation programs have been recommended [27,28]. The STST has been used to examine healthy adults but also patients with lung disease, renal disease, stroke, osteoporosis or patients in palliative care [26]. Direct comparisons of the STST and the 6-minute walk test in different settings showed differences in cardiorespiratory demand but comparable estimates of functional exercise capacity [27,29,30]. However, although the 6-minute walk test and step tests are well-established field tests, they were primarily developed to assess functional exercise capacity or cardiorespiratory fitness rather than to provide a standardized exercise stimulus for investigating exercise-induced immune regulation [26–30]. Moreover, the physiological stimulus elicited by these tests is determined by different methodological characteristics. Performance during the 6-minute walk test is influenced by its self-paced nature and a documented learning effect associated with repeated testing [31,32], whereas the Chester Step Test is an incremental exercise test in which exercise duration and the achieved workload depend on the participant’s performance and termination point [33,34]. Consequently, participants are exposed to different cumulative exercise stimuli despite a standardized test protocol, which may complicate the use of the test as a standardized experimental model for eliciting comparable acute immune-metabolic responses. By contrast, the 1-minute STST consists of a brief, fixed-duration maximal-effort task that minimizes the influence of prolonged pacing strategies while providing a near-maximal stimulus. This may offer a more standardized exercise challenge for investigating acute immune-metabolic responses. The present study therefore investigates whether the STST is capable of eliciting sufficient metabolic stress, reflected by lactate accumulation above the anaerobic threshold, along with characteristic immune cell redistribution. Taken together, these characteristics suggest that the STST may represent a practical field-based alternative for standardized immune-metabolic assessment in settings where laboratory-based exercise testing is impractical or unavailable.

Therefore, this study aimed not only to characterize the metabolic and immunological responses to the STST, but also to evaluate whether this simple field test can serve as a standardized model for investigating exercise-induced immune-metabolic stress responses outside laboratory settings. We hypothesize that the STST will induce blood lactate concentrations of more than 4 mmol/L and is associated with measurable exercise-induced changes in immune cell counts (white blood cell count, lymphocytes, granulocytes and associated immune-inflammatory indices).

## 2 Methods

### 2.1 Participants and study design

All participants were apparently healthy and uninjured (i.e., no acute symptoms or pain). Participants were sedentary and recreationally active according to McKay *et al.* [35], and exhibited lightly to moderately irregular physical activity patterns. The sample size was determined based on previous studies investigating acute exercise-induced alterations in circulating immune markers in healthy adults [22,36–38]. Therefore, a sample size of 23 participants was considered appropriate to detect acute exercise-induced changes in immune and metabolic parameters using mixed linear model regression analyses. To account for possible dropouts, 28 participants were recruited. The test was carried out after an overnight fast, during which no food or fluids other than water were consumed. All participants were advised to avoid physical activity right before the test and intensive exercise on the day before the measurement. Before the test began, the participants’ age, height and weight were recorded. The test started with a 30-minute resting phase in a sitting position in order to be able to depict realistic resting values (Figure 1). During the resting phase, capillary blood was taken from the earlobe after 10 and 20 minutes for blood count, lactate and glucose analysis. After the initial resting phase, the 1-minute STST was performed (protocol described in the next section). The test was followed by a second resting phase of 45 minutes in a sitting position. As in the first rest period, participants were free to spend the second resting phase as they liked, as long as it was not physically or mentally demanding and they remained seated (e.g. looking at phones, talking to experimental staff). Blood samples were taken immediately after the test and 5, 10, 15, 30, and 45 minutes post-exercise. Heart rate and oxygen saturation were recorded parallel to the blood sampling times and additionally every minute in the first 5 minutes after the exercise test with a pulse oximeter (*PC-60B1 Fingertip, Creative Industry Co.*). All participants were informed about the type and procedure of the test before they gave their written consent. This study was carried out in accordance with the recommendations of the University Research Ethics Committee of the Friedrich Schiller University Jena, Germany and the latest version of the Declaration of Helsinki. The Protocol was approved by the University Ethics Committee of the Friedrich Schiller University/University Hospital Jena (458510/15, 2025-3763-BO-A).

**Figure 1.**
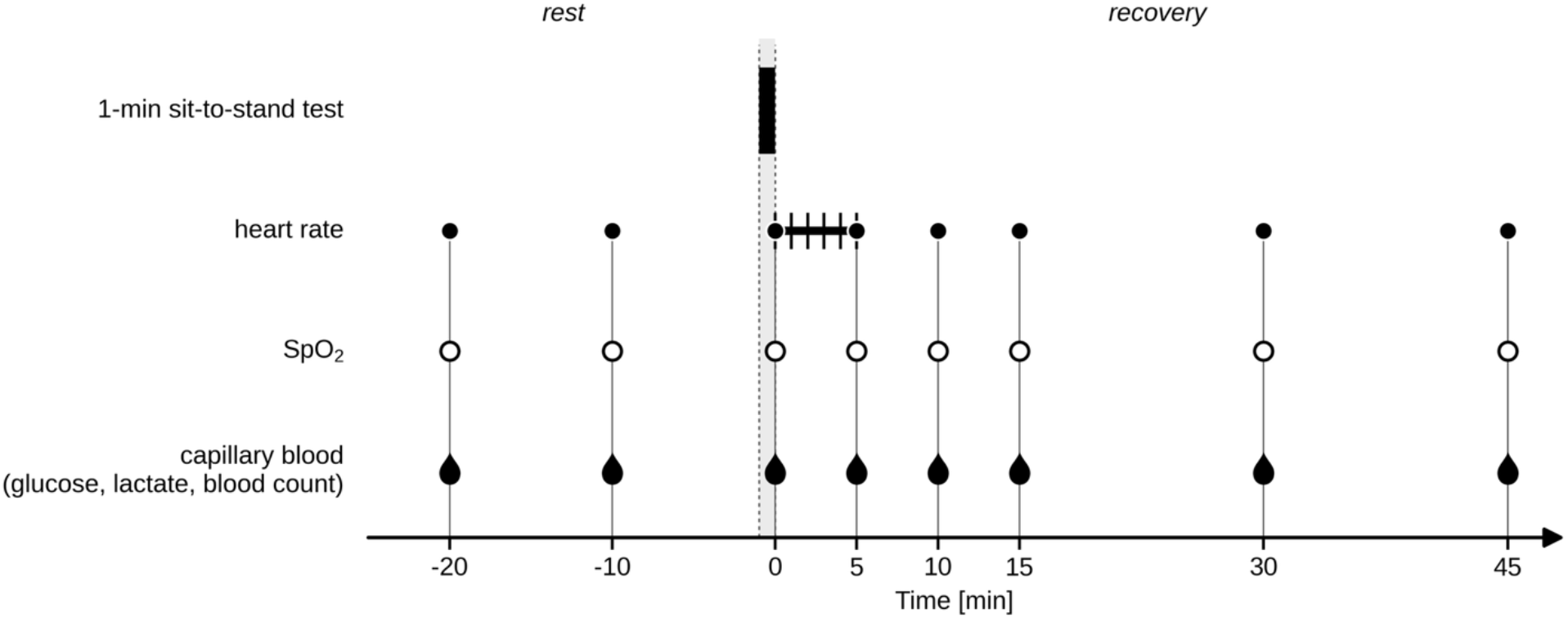
Study design. Timing of the 1-minute sit-to-stand test (black bar), capillary blood sampling for glucose, lactate, and blood counts (drops), heart rate and oxygen saturation (SpO2) recording.

### 2.2 1-minute sit-to-stand test (STST)

The 1-minute STST is a simple exercise test that can be used to assess exercise tolerance and physical performance with minimal material and time expenditure. The test was carried out according to Ozalevli *et al.* [29]. The participants had to repeatedly stand up from a chair and sit down again, whereby briefly touching the chair was sufficient before standing up again as quickly as possible within one minute. The legs should be fully extended when standing and bent at about 90° when sitting. During the entire test the legs should be about shoulder-width apart and the arms crossed in front of the chest. The chair used for the test had a standard height, no armrests and was placed against a stable desk to avoid any movement of it. All participants used the same chair. The number of complete sit-to-stand cycles was counted. The participants had the opportunity to perform a trial test of one cycle right before starting the test.

### 2.3 Blood count and lactate/glucose measurement

At the beginning, the earlobe of the study participants was rubbed with Finalgon® ointment to promote blood circulation. The ointment was removed from the ear using an alcohol-soaked swab before it was punctured with a lancet for blood collection. Finalgon® was used solely to facilitate capillary blood sampling. The active ingredients, nonivamide and nicoboxil, exert their effects locally by stimulating afferent nerve endings in the skin and inducing vasodilation, respectively [39]. Based on its established mechanism of action and available safety data, topical nonivamide/nicoboxil has not been reported to induce systemic immunomodulatory effects. However, local preanalytical effects resulting from increased skin perfusion on the composition of capillary blood cannot be completely excluded. But since the ointment was applied to all study participants according to a standardized procedure (identical application dose, exposure time, and sampling site), any potential local effects represent a consistent component of the sampling procedure and are therefore unlikely to introduce systematic bias into comparisons within the study. For each capillary blood collection, a safety lancet was inserted into the earlobe of the participant and 20 µL of capillary blood was taken into an EDTA-coated plastic capillary for blood cell count analysis and 20 µL were taken into a glass capillary for lactate and glucose analysis. The blood cell count analysis was performed immediately after collection using a hematology analyser (*Medonic M-series M32M, Boule Medical AB*). For the determination of lactate and glucose, the samples from all measurement times were analyzed after the last blood draw using enzymatic-amperometric measurement (*Super GL speedy, Dr. Müller Gerätebau GmbH*). The samples taken in the capillary were each placed in a reaction vessel filled with a defined quantity (1000 µL) of hemolysis system solution immediately after collection and placed in a sample reservoir in the device. In accordance with the manufacturer’s instructions, the time between sample collection and dilution with the hemolysis solution did not exceed 15 minutes, and hemolyzed samples were stored at temperatures no more than 23°C for a maximum of 80 minutes before analysis (manufacturer’s limit: up to 12 hours at a temperature below 23°C).

Analyzed blood biomarkers were leukocyte count (white blood cell count, WBC), lymphocyte count (LYM), granulocyte count (GRA), mid-sized white cells (MID), and platelet count (PLT). Changes in plasma and blood volume after exercise were calculated using the Dill and Costill equation [40] and blood biomarkers were corrected according to Matomäki *et al.* [41]. The results are given as uncorrected und corrected values. Corrected values were used to calculate the neutrophil-to-lymphocyte ratio (NLR), Systemic immune-inflammation index (SII) and systemic inflammatory response index (SIRI) [42]. As differential neutrophil counts were not available due to capillary blood measurement, GRA were used as a surrogate for neutrophil counts to calculate NLR, SII and SIRI. Given that neutrophils constitute the vast majority of circulating granulocytes under physiological conditions, this approach provides a reasonable approximation of these indices. According to this approximation, the term GLR (granulocyte-to-lymphocyte ratio) was used instead of NLR. SII and SIRI were calculated as follows:

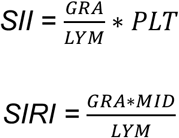

### 2.4 Statistical analysis

The data was analyzed using *Microsoft Excel* software version 2406 and the statistical analyses were performed with *Python* version 3.12.7 and *Jupyter Notebook* version 7.2.2. Potential outliers were identified using the Grubbs test. Statistical outliers were excluded only if they were considered attributable to measurement error or to factors unrelated to the physiological response to exercise. Comparisons in baseline characteristics between men and women were conducted using Welch’s t-test. Mixed linear Model Regression analyses were performed to determine differences between measurement times and sex. Normality of the data was tested using the Shapiro-Wilk test. Pearson’s and Spearman’s correlation analyses were conducted to assess relationships between variables. Pearson’s correlation coefficient (r) was used to evaluate linear relationships between continuous variables. In addition, Spearman’s rank correlation coefficient (ρ) was calculated to assess monotonic relationships. For both correlation measures, the corresponding correlation coefficients are reported. A p-value < 0.05 was considered statistically significant. The results are given as mean ± standard deviation.

## 3 Results

To investigate the acute normative immunological stress regulations that occur in response to a single bout of exercise, a clinical study that included 28 healthy participants (17 women and 11 men, aged 23.70 ± 2.00 years, with a BMI of 23.39 ± 2.16 kg/m^2^) was performed. The study was conducted from July 2023 to February 2024 in Jena, Germany. First, the baseline characteristics of the participants included in the analysis were evaluated; however, height and weight were recorded only after the first 6 participants (Table 1). Except for height and weight (p < 0.05 for both) no differences in baseline characteristics were observed between men and women. The maximum heart rate (HRmax) of the participants was calculated according to Nes *et al.* [43].

**Table 1.**
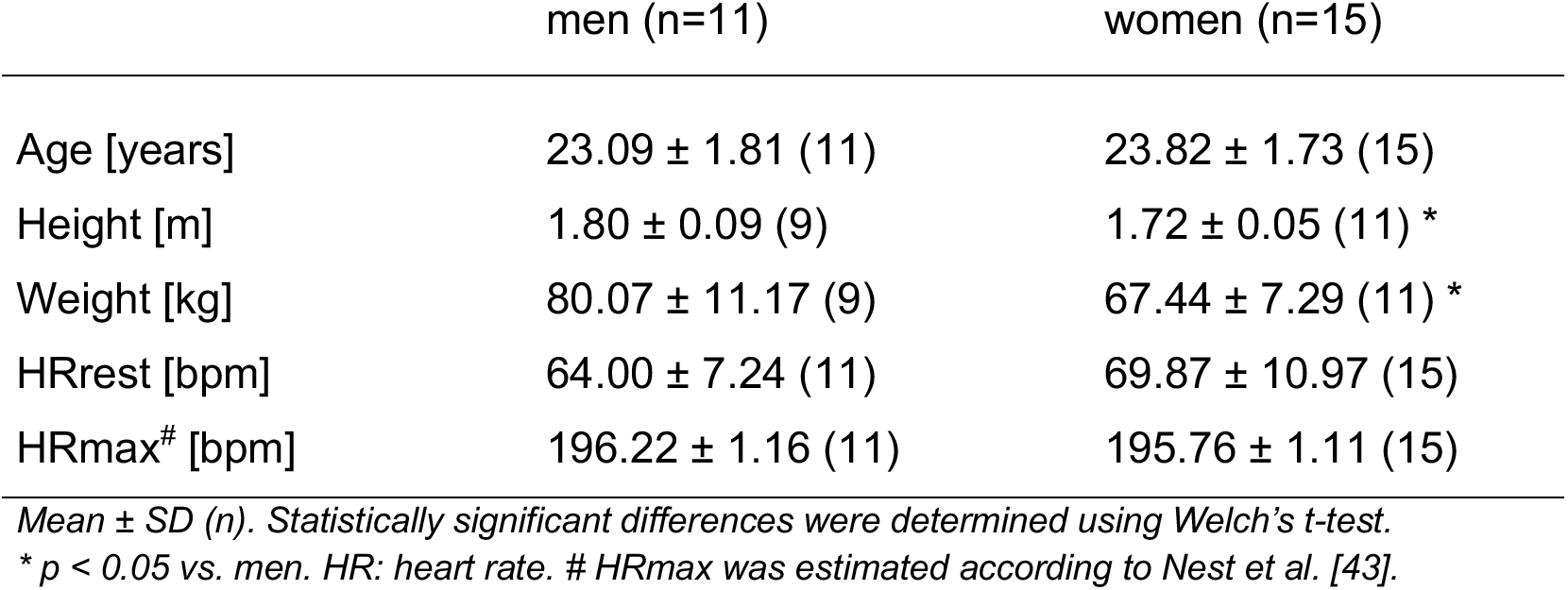
Characteristics of the participants.

To investigate cardiorespiratory, metabolic, immunological, and inflammatory biomarkers, blood sampling as well as heart rate and oxygen saturation measurements were performed before and after the STST (Table 2). During the test, the participants completed an average of 56 sit-to-stand cycles (min: 44 cycles; max: 87 cycles). No sex differences were observed for the number of cycles. One of the 28 participants was excluded from the analyses because multiple independent immune parameters were identified as statistical outlier values by the Grubbs test and were considered unlikely to reflect the individual’s physiological response to exercise. Another participant was excluded due to measurement error, resulting in missing values. The participants’ oxygen saturation fell immediately after the STST below the baseline level obtained 10 minutes before the test (p < 0.01) and differed between women and men at every timepoint with women showing higher values than men (p < 0.01). The heart rate increased significantly immediately after the test to an average peak heart rate of 132.19 ± 18.59 bpm (p < 0.001) and remained significantly elevated until 15 minutes after the test (75.81 ± 9.51 bpm) compared to the resting heart rate (-10 min) of 67.38 ± 9.86 bpm (p < 0.01). To draw conclusions about the training status of the participants, the heart rate recovery was also calculated as the difference in heart rate between the maximum heart rate (0 min) and the measured heart rate in minutes 1 to 5 after the STST. As the heart rate measurement per minute in the first 5 minutes after the test was introduced after the first 6 participants, the heart rate recovery data is only available from 20 of the 26 participants. In the first minute after the exercise, the heart rate recovery was on average 22.65 ± 12.88 bpm (min: 4 bpm; max: 51 bpm). After 5 minutes the heart rate dropped by 50.65 ± 13.43 bpm (min: 27 bpm; max: 86 bpm).

**Table 2.** Results for biomarker changes before (-20, -10) and after the 1-minute STST (0, 1, 2, 3, 4, 5, 10, 15, 30, 45) in capillary blood of the participants (n=26).

| time point [min] | -20 | -10 | 0 | 1 | 2 | 3 | 4 | 5 | 10 | 15 | 30 | 45 |
| --- | --- | --- | --- | --- | --- | --- | --- | --- | --- | --- | --- | --- |
| **Cardiorespiratory** |  |  |  |  |  |  |  |  |  |  |  |  |
| HR [bpm] | 69.88 ± 9.14<br>### | 67.38 ± 9.86<br>### | 132.19 ± 18.59 *** | 107.75 ± 24.33 *** , ### | 89.00 ± 23.04<br>*** , ### | 81.65 ± 18.18<br>*** , ### | 80.40 ± 16.74<br>*** , ### | 80.69 ± 13.23<br>*** , ### | 77.08 ± 11.20<br>*** , ### | 75.81 ± 9.51<br>** , ### | 72.92 ± 8.60<br>### | 69.42 ± 8.70<br>### |
| HRR [bpm]<br>(n=20) | - | - | - | 22.65 ± 12.88 | 41.40 ± 11.89 | 48.75 ± 11.10 | 50.00 ± 11.72 | 50.65 ± 13.43 | - | - | - | - |
| oxygen saturation [%] | 98.23 ± 0.95<br>### | 98.08 ± 0.98<br>### | 97.15 ± 2.51<br>*** | 97.80 ± 0.89<br>### | 97.95 ± 0.76<br>### | 98.00 ± 0.65<br>### | 97.65 ± 0.81<br>### | 97.81 ± 0.85 ## | 97.77 ± 0.91 ## | 97.62 ± 1.06 # | 97.54 ± 1.14 *, # | 97.85 ± 0.83<br>### |
| **Metabolic** |  |  |  |  |  |  |  |  |  |  |  |  |
| lactate [mmol/L] | 0.79 ± 0.24 ### | 0.74 ± 0.21 ### | 4.63 ± 1.70 *** | - | - | - | - | 5.83 ± 1.92<br>*** , ### | 4.74 ± 1.78 *** | 3.77 ± 1.49<br>*** , ### | 2.02 ± 0.75<br>*** , ### | 1.39 ± 0.51 **, ### |
| lactate [mmol/L],<br>corrected | 0.79 ± 0.24 ### | 0.74 ± 0.21 ### | 4.50 ± 1.66 *** | - | - | - | - | 5.76 ± 1.87<br>*** , ### | 4.72 ± 1.76 *** | 3.77 ± 1.49<br>*** , ## | 2.03 ± 0.78<br>*** , ### | 1.43 ± 0.58 **, ### |
| glucose [mmol/L] | 5.06 ± 0.60 **, ### | 5.34 ± 0.53 | 5.51 ± 0.62 | - | - | - | - | 5.45 ± 0.60 | 5.12 ± 0.65 *, ### | 5.24 ± 0.45 ## | 4.97 ± 0.39<br>*** , ### | 4.90 ± 0.33<br>*** , ### |
| glucose [mmol/L],<br>corrected | 5.06 ± 0.60 *, # | 5.34 ± 0.53 | 5.36 ± 0.70 | - | - | - | - | 5.38 ± 0.57 | 5.09 ± 0.71 *, # | 5.25 ± 0.52 | 4.99 ± 0.45 **, ## | 5.00 ± 0.43 **, ## |
| **Immunological** |  |  |  |  |  |  |  |  |  |  |  |  |
| WBC [10<sup>9</sup>/L] | 6.37 ± 1.42 **, ### | 5.55 ± 1.28 ### | 8.41 ± 2.21 *** | - | - | - | - | 7.35 ± 2.08<br>*** , ### | 6.72 ± 2.18<br>*** , ### | 6.25 ± 1.80 **, ### | 5.70 ± 1.50 ### | 6.04 ± 1.75 ### |
| WBC [10<sup>9</sup>/L],<br>corrected | 6.37 ± 1.42<br>*** , ### | 5.55 ± 1.28 ### | 8.10 ± 1.88 *** | - | - | - | - | 7.25 ± 2.01<br>*** , ### | 6.67 ± 2.16<br>*** , ### | 6.24 ± 1.76 **, ### | 5.74 ± 1.57 ### | 6.18 ± 1.94 **, ### |
| LYM [10<sup>9</sup>/L] | 2.62 ± 0.59 **, ### | 2.23 ± 0.47 ### | 3.97 ± 1.03 *** | - | - | - | - | 3.03 ± 0.77<br>*** , ### | 2.63 ± 0.93 **, ### | 2.32 ± 0.71 ### | 2.02 ± 0.57 ### | 2.04 ± 0.68 ### |
| LYM [10<sup>9</sup>/L],<br>corrected | 2.62 ± 0.59 **, ### | 2.23 ± 0.47 ### | 3.82 ± 0.87 *** | - | - | - | - | 2.99 ± 0.73<br>*** , ### | 2.61 ± 0.90 **, ### | 2.31 ± 0.64 ### | 2.03 ± 0.57 ### | 2.07 ± 0.64 ### |
| GRA [10<sup>9</sup>/L] | 3.29 ± 1.06 *, ### | 2.92 ± 0.94 ### | 3.82 ± 1.43 *** | - | - | - | - | 3.78 ± 1.41 *** | 3.57 ± 1.31 *** | 3.46 ± 1.19<br>*** , # | 3.26 ± 1.04 *, ### | 3.51 ± 1.30<br>*** , # |
| GRA [10<sup>9</sup>/L],<br>corrected | 3.29 ± 1.06 *, ## | 2.92 ± 0.94 ### | 3.68 ± 1.32 *** | - | - | - | - | 3.73 ± 1.37 *** | 3.55 ± 1.32 *** | 3.46 ± 1.23 *** | 3.29 ± 1.10 *, ## | 3.61 ± 1.49 *** |
| MID [10<sup>9</sup>/L] | 0.47 ± 0.14 *, ### | 0.40 ± 0.13 ### | 0.62 ± 0.18 *** | - | - | - | - | 0.54 ± 0.18<br>*** , ## | 0.52 ± 0.21<br>*** , ## | 0.47 ± 0.17 *, ### | 0.43 ± 0.16 ### | 0.48 ± 0.16 **, ### |
| MID [10<sup>9</sup>/L],<br>corrected | 0.47 ± 0.14 *, ### | 0.40 ± 0.13 ### | 0.60 ± 0.15 *** | - | - | - | - | 0.53 ± 0.17<br>*** , # | 0.52 ± 0.20<br>*** , ## | 0.47 ± 0.17 *, ### | 0.43 ± 0.15 ### | 0.49 ± 0.17 **, ### |
| PLT [10<sup>9</sup>/L] | 170.38 ± 30.42 | 158.92 ± 34.32 | 163.50 ± 39.87 | - | - | - | - | 156.31 ± 34.12 | 160.42 ± 42.35 | 151.77 ± 48.60 | 167.31 ± 32.36 | 152.96 ± 37.00 |
| PLT [10<sup>9</sup>/L],<br>corrected | 170.38 ± 30.42 | 158.92 ± 34.32 | 157.61 ± 37.18 | - | - | - | - | 154.73 ± 35.12 | 159.17 ± 41.92 | 150.58 ± 46.00 | 167.73 ± 32.28 | 155.34 ± 35.87 |
| **Inflammatory** |  |  |  |  |  |  |  |  |  |  |  |  |
| GLR | 1.30 ± 0.45 ### | 1.33 ± 0.40 ### | 1.00 ± 0.38 *** | - | - | - | - | 1.27 ± 0.38 ### | 1.41 ± 0.44 ### | 1.53 ± 0.47 **, ### | 1.67 ± 0.54<br>*** , ### | 1.80 ± 0.67<br>*** , ### |
| SII | 223.01 ± 88.71 ### | 216.51 ± 87.64 ## | 158.36 ± 73.86 ** | - | - | - | - | 198.08 ± 75.71 # | 223.38 ± 85.63 ### | 234.71 ± 120.95 ### | 282.59 ± 120.31 *** , ### | 286.57 ± 143.92 *** , ### |
| SIRI | 0.62 ± 0.32 | 0.55 ± 0.30 | 0.61 ± 0.32 | - | - | - | - | 0.71 ± 0.42 * | 0.74 ± 0.44 **, # | 0.75 ± 0.44 **, # | 0.72 ± 0.34 **, # | 0.93 ± 0.59<br>*** , ### |

Mean ± SD. Statistically significant differences were determined using Mixed linear Model Regression and are shown for the measurement times -10 min and 0 min as \* $p < 0.05$ , \*\* $p < 0.01$ , \*\*\* $p < 0.001$ vs. -10 min and # $p < 0.05$ , ## $p < 0.01$ , ### $p < 0.001$ vs. 0 min. GLR: granulocyte-to-lymphocyte ratio, GRA: granulocyte count, HR: heart rate, HRR: heart rate recovery, LYM: lymphocyte count, MID: mid-sized white cells, PLT: platelet count, SII: Systemic immune-inflammation index, SIRI: systemic inflammatory response index, WBC: white blood cell count.

Changes in blood volume, especially plasma volume, were observed (Figure 2) and accordingly the obtained values of blood biomarkers were corrected for hemoconcentration. Only corrected values are discussed hereafter, while uncorrected values are provided in Table 2.

**Figure 2.**
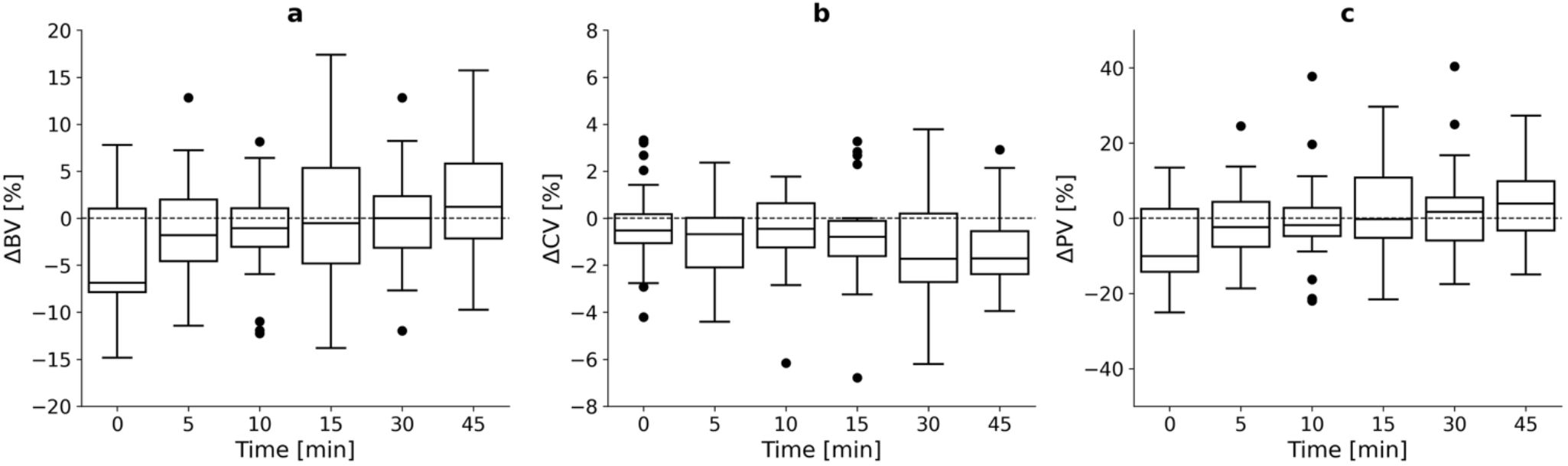
Volume changes of blood volume (ΔBV) (a), red cell volume (ΔCV) (b) and plasma volume (ΔPV) (c) after the 1-minute STST.

Next, to objectively assess the intensity of the STST, glucose and lactate concentrations were recorded during the test. Analysis showed that the glucose concentration was significantly increased immediately after the STST compared to the beginning of the test (-20 min) (5.06 ± 0.60 mmol/L vs. 5.36 ± 0.70 mmol/L) (p < 0.05) but did not change compared to 10 minutes before. After 10 minutes post-exercise the concentration fell below the values measured directly after and 10 minutes before exercise (p < 0.05 for both). Glucose concentration significantly decreased again 30 minutes post-exercise (p < 0.01 vs. -10 min) (Figure 3 a). Further, lactate concentration increased immediately after exercise (p < 0.001). With 5.76 ± 1.87 mmol/L, the maximum concentration was usually reached 5 minutes after the test, which depicts an almost 7-fold increase compared to the resting lactate value of 0.74 ± 0.21 mmol/L (-10 min) (p < 0.001). Until the end of the measurements (45 min), the lactate concentration remained with 1.43 ± 0.58 mmol/L significantly elevated compared to the resting lactate value (p < 0.01) (Figure 3 a).

**Figure 3.**
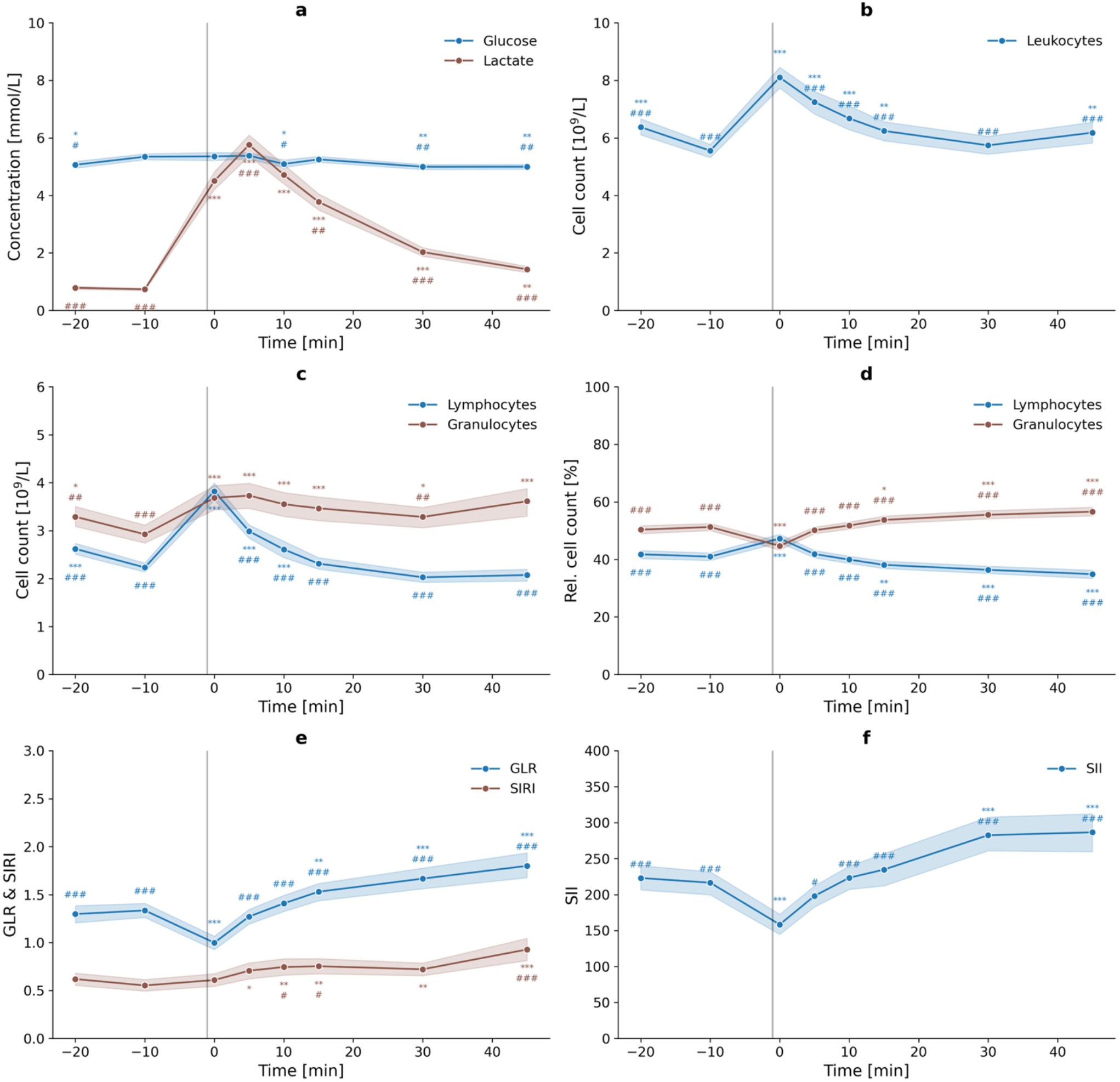
Glucose and lactate (a), leukocyte count (b), lymphocyte and granulocyte count absolute (c) and relative (d) measured in capillary blood before and after the 1-minute STST. Granulocyte-to-lymphocyte ratio (GLR), and systemic inflammatory response index (SIRI) (e) and Systemic immune-inflammation index (SII) (f) were calculated based on the obtained biomarker values corrected for hemoconcentration. The vertical line marks the time of the test. Mean and error band are shown per time point. Statistically significant differences were determined using Mixed linear Model Regression and are shown for the measurement times -10 min and 0 min separated by parameter (above or below and colored according to the parameter). * p < 0.05, ** p < 0.01, *** p < 0.001 vs. -10 min and # p < 0.05, ## p < 0.01, ### p < 0.001 vs. 0 min.

To examine the exercise-induced immunological stress response, total WBC (Figure 3 b) and cell counts of the WBC subpopulations (Figure 3 c) were determined. During the 30-minute resting phase before the STST, there was initially a significant decrease (p < 0.001) in WBC of about 13 %, whereby there was no difference between women and men. The data showed that immediately after the STST, total WBC significantly increased in both sexes by 46 % compared to the resting value 10 minutes before the test (p < 0.001). Total WBC decreased gradually after the test until the initial value was almost reached again after 30 minutes. At no time point after the exercise did WBC differ between the sexes.

The changes in total WBC observed during the experiment were also reflected in its subpopulations. In the pre-exercise phase, LYM decreased significantly by 15 % between the first and second measurement time point (p < 0.01). The GRA decreased by 11 % (p < 0.05). Immediately after the STST, the cell counts were significantly increased compared to the resting value (-10 min), whereby LYM increased by 71 % (p < 0.001) and GRA increased by 26 % (p < 0.001). Post-exercise LYM values decreased and tend to fall below the resting level after 30 minutes (p = 0.07). The GRA decreased slightly after 5 minutes until 30 minutes post-exercise but did not return to baseline levels, instead slightly increased again 45 minutes post-exercise.

The percentages of LYM and GRA in the leukocytes showed an opposing dynamic (Figure 3 d). The proportion of LYM significantly increased immediately after the test compared to the proportion 10 minutes before the STST (from 40.95 ± 6.49 % to 47.26 ± 7.51 %), while the proportion of GRA significantly decreased (from 51.25 ± 6.53 % to 44.68 ± 7.62 %). Both proportions thus differed immediately post-exercise significantly from the resting value 10 minutes before the test (p < 0.001 for both). Immediately post-exercise the population proportions reversed and the LYM proportion decreased until the end of the measurement (45 min), while the GRA proportion increased. Both percentages differed 30 and 45 minutes post-exercise significantly from those measured before (-10 min) (p < 0.001 for all) and immediately after exercise (0 min) (p < 0.001 for all). It was also shown for both leukocyte populations that the respective changes in the proportions were initially greater in the post-exercise phase and were lower from 15 minutes until the end of post-exercise measurements. Thus, the respective initial level was reached again between 5 and 10 minutes after the STST and subsequently decreased below (LYM) or increased above (GRA) the resting value before the exercise test.

Next, to estimate the inflammatory status of the participants, the GLR, SII and SIRI were calculated (Figure 3 e and f). The GLR significantly decreased immediately after the STST compared to 10 minutes before (p < 0.001), returned to baseline level after 5 minutes and increased after that until 45 minutes post-exercise (p < 0.001 for 30 and 45 min). The SII was slightly lower for men than for women during the entire measurement period but did not differ significantly. Immediately after the STST, the SII was significantly decreased compared to 20 (p < 0.001) and 10 minutes before (p < 0.01), regardless of sex. The SII then increased until 30 minutes after the STST the baseline level (-10 min) was exceeded by 31 % (p < 0.001). Compared to the average SII immediately post-exercise, the value increased by 78 % (p < 0.001). The SIRI remained unchanged immediately after the exercise but rose post-exercise and was significantly increased at 5 minutes after the test compared to 10 minutes before (p < 0.05). Until 45 minutes post-exercise the SIRI continued to rise and was significantly elevated at this timepoint compared to 10 minutes before (p < 0.001) and immediately after (p < 0.001) the test.

Moreover, to investigate the influence of exercise intensity and/or training status on the magnitude of biomarker changes following exercise, correlation analyses were conducted. Changes in heart rate from baseline to 5 minutes post-exercise (ΔHR) ranged from -1 to 33 bpm and correlated significantly with the corresponding changes in lactate (ΔLactate; r = 0.72, ρ = 0.62, both p < 0.001), leukocyte count (ΔWBC; r = 0.68, ρ = 0.64, both p < 0.001) and lymphocyte count (ΔLYM; r = 0.64, ρ = 0.64, both p < 0.001) (Figure 4). Changes in granulocyte count (ΔGRA) showed a significant Pearson correlation with ΔHR (r = 0.53, p < 0.01), whereas Spearman correlation indicated a trend (ρ = 0.36, p = 0.07). However, heart rate recovery 5 minutes after the exercise test was not correlated with any of the metabolic, immunological, or inflammatory markers at that time point.

**Figure 4.**
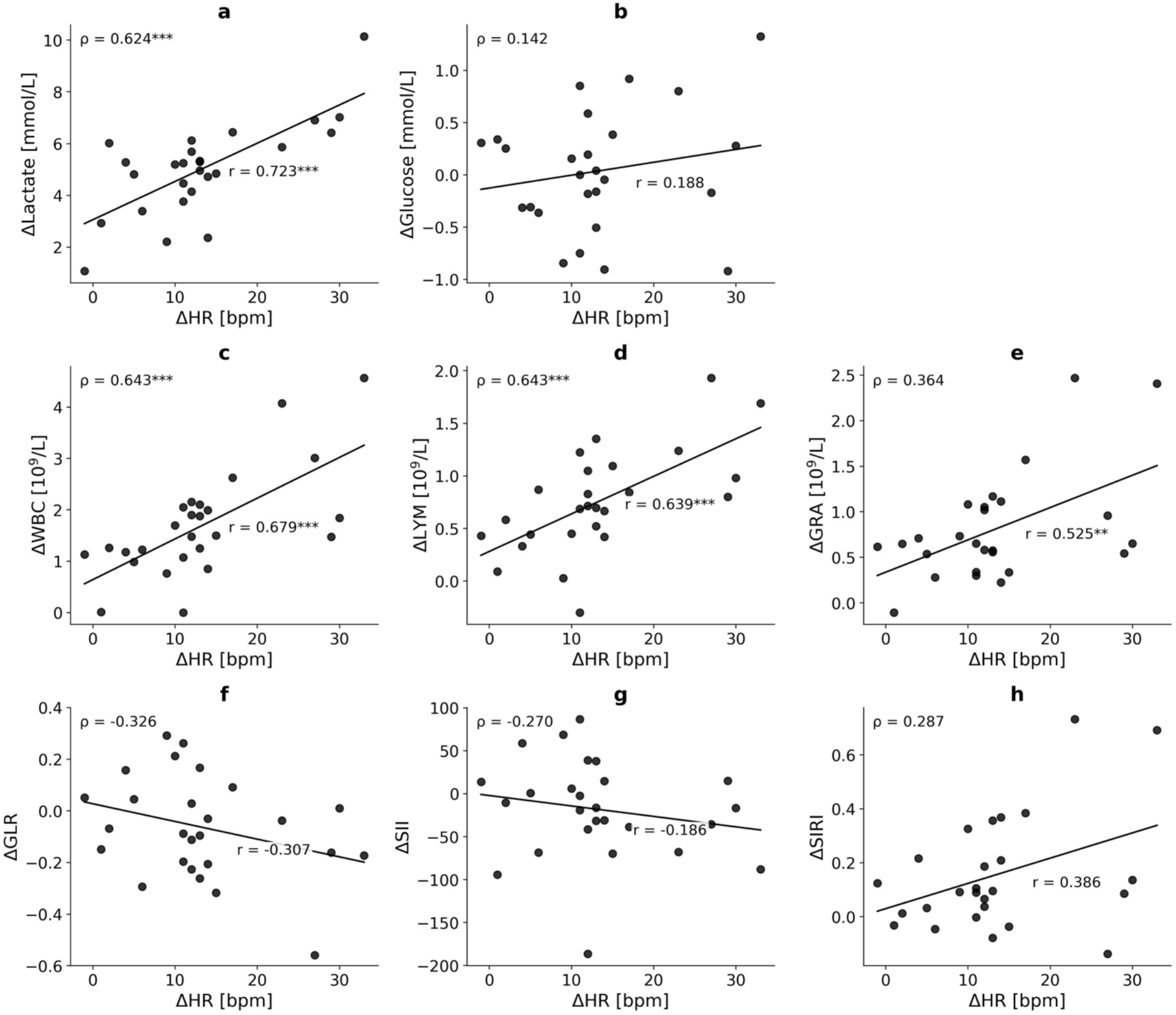
Correlations of changes in heart rate from baseline to 5 minutes post-exercise (ΔHR) and corresponding changes in lactate concentration (ΔLactate) (a), glucose concentration (ΔGlucose) (b), white blood cell count (ΔWBC) (c), lymphocyte count (ΔLYM) (d), granulocyte count (ΔGRA) (e), granulocyte-to-lymphocyte ratio (ΔGLR) (f), systemic immune-inflammation index (ΔSII) (g) and systemic inflammatory response index (ΔSIRI) (h). GLR, SII and SIRI were calculated based on the obtained biomarker values corrected for hemoconcentration. Pearson’s (r) and Spearman’s (ρ) correlation coefficients are shown for each panel. ** p < 0.01, *** p < 0.001.

Lastly, to further examine the relationship between metabolic and immune responses to exercise, correlations between changes in lactate concentration and immune parameters were assessed. Changes in lactate concentration from baseline to 5 minutes post-exercise (ΔLactate) ranged from 1.07 to 10.14 mmol/L and correlated significantly with corresponding ΔWBC (r = 0.71, ρ = 0.65, both p < 0.001), ΔLYM (r = 0.64, ρ = 0.58, p < 0.01 and p < 0.001) and ΔGRA (r = 0.60, ρ = 0.52, both p < 0.01) (Figure 5).

**Figure 5.**
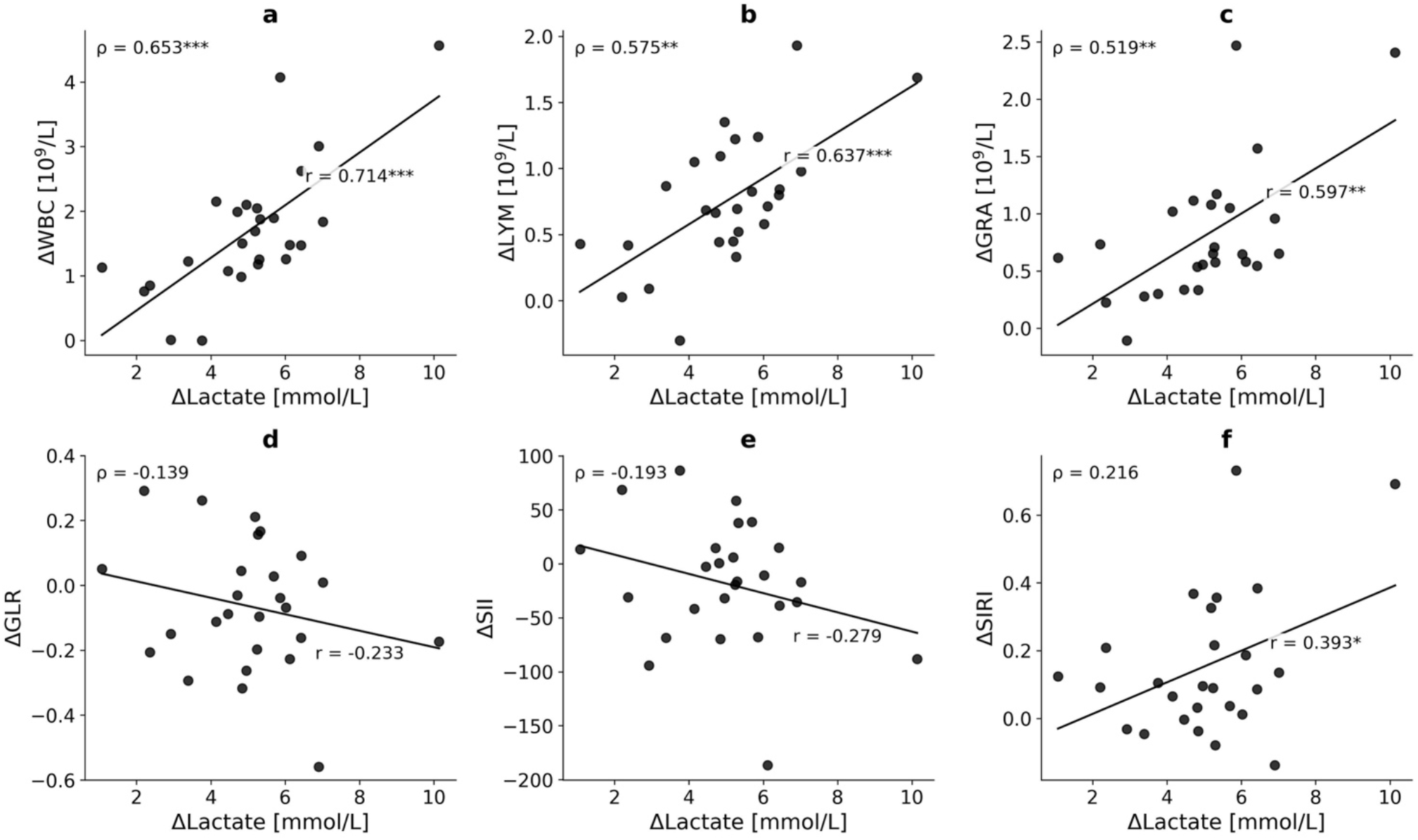
Correlations of changes in lactate concentration from baseline to 5 minutes post-exercise (ΔLactate) and corresponding changes in white blood cell count (ΔWBC) (a), lymphocyte count (ΔLYM) (b), granulocyte count (ΔGRA) (c), granulocyte-to-lymphocyte ratio (ΔGLR) (d), systemic immune-inflammation index (ΔSII) (e) and systemic inflammatory response index (ΔSIRI) (f). GLR, SII and SIRI were calculated based on the obtained biomarker values corrected for hemoconcentration. Pearson’s (r) and Spearman’s (ρ) correlation coefficients are shown for each panel. * p < 0.05, ** p < 0.01, *** p < 0.001.

## 4 Discussion

The 1-minute STST is an established performance test in the investigation of various diseases [44]. The aim of this study was to examine the effects of the 1-minute STST on immune-metabolic stress indices and to determine whether it elicits a sufficiently high intensity to qualify as an anaerobic exercise test, thereby supporting its application for investigating exercise-induced immunological stress regulation. With an average of 56 complete sit-to-stand cycles in 1 minute, the participants demonstrated high functional performance. Compared with age- and sex-specific reference values, their performance corresponded to the 75^th^ percentile according to Strassmann *et al.* [45] and to the 75^th^–97.5^th^ percentile according to Otto-Yáñez *et al.* [46], while completing considerably more cycles than those reported by Furlanetto *et al.* [47] and Vilarinho *et al.* [44]. Several studies have reported that the 1-minute STST demonstrates high test–retest reliability [26]. However, it can be assumed that correct implementation of the test, participant motivation, and individual participant attributes such as height and weight (which showed no correlation with the number of completed sit-to-stand repetitions in the present study) may have a considerable effect on the test performance. Therefore, to draw more reliable conclusions regarding test intensity or the participants’ training status, heart rate data including percentage of the individual maximum heart rate and heart rate recovery were additionally considered.

In addition to muscle strain, acute intensive exercise is accompanied by an increase in heart rate during to immediately after exercise due to the release of catecholamines [11]. Consistently, a significant increase in heart rate from a resting heart rate of 67.38 ± 9.86 bpm to a peak heart rate of 132.19 ± 18.59 bpm was observed immediately after the test. Compared to the heart rate measured by Ozalevli *et al.* at the end of the STST in healthy 63 ± 8 years old individuals and COPD-patients the average peak heart rate of the participants was considerably higher [29]. However, also number of completed sit-to-stand cycles exceeded those reported by Ozalevli *et al.* notably. The peak heart rate corresponds to 67.47 ± 9.90 % of the estimated maximum heart rate of the participants (195.95 ± 1.13 bpm). According to Garber *et al.* this represents a moderate intensity of the exercise test [48]. However, heart rate measurements immediately after exercise using a fingertip pulse oximeter may be affected by measurement delay and reduced accuracy during rapid changes in heart rate, which could result in an underestimation of the peak heart rate values.

Previous work has indicated that heart rate recovery after exercise is related to training status and aerobic fitness, with higher differences between maximum heart rate and post-exercise heart rate typically observed in trained compared with untrained individuals [49]. The average heart rate recovery in the first minute after the STST was 22.65 ± 12.88 bpm, which is comparable to the heart rate recovery calculated by Römer and Wolfarth 1 minute after cycle ergometer exercise tests [50]. However, the range of heart rate recovery after 1 minute between participants in the present study was high (min: 4 bpm; max: 51 bpm). Thereby the participant with the lowest heart rate recovery at 1 minute post-exercise (4 bpm) had a resting heart rate of 80 bpm and a maximum heart rate of 120 bpm, while the participant with the highest heart rate recovery (51 bpm) had a resting heart rate of 53 bpm and a maximum heart rate of 114 bpm. Up to 5 minutes after the test, the heart rate had decreased on average by 50.65 ± 13.43 bpm, which is about 10 bpm below the heart rate recovery determined by Römer and Wolfarth [50]. Even 5 minutes after the STST, the heart rate recovery varied greatly between individuals, with a maximum recovery of 86 bpm (participant had a resting heart rate of 55 bpm and maximum heart rate of 164 bpm) and a minimum recovery of 27 bpm (resting heart rate: 80 bpm; maximum heart rate: 120 bpm). The striking differences in the heart rate recovery may indicate differences in the training status of the participants. However, no universal threshold values for heart rate recovery have been established to classify training status, as heart rate recovery is strongly affected by other factors such as the exercise protocol, individual characteristics, and recovery conditions [49,50]. In the present study, a smaller post-exercise decrease in heart rate could also reflect that the test provoked only a modest increase in heart rate, i.e., a smaller difference between maximum and resting heart rate, as observed in several participants. For participants with both a smaller heart rate recovery and a rather low number of cycles, it can be assumed that the test intensity was low due to the implementation and motivation (particularly reflected in a lower pacing), resulting in a small change in heart rate and heart rate recovery.

As intensive exercise is associated with increasing lactate production, the intensity of the STST was also assessed by the change in lactate concentrations and glucose levels due to the exercise. Furthermore, the anaerobic threshold was used as a physiologically relevant reference point, as it marks the threshold beyond which metabolic and neuroendocrine responses increase substantially [51]. Exercise-induced immune responses are closely linked to cellular metabolism, as immune cell activation and function are strongly influenced by metabolic signals and substrate availability [24,52]. Beyond the anaerobic threshold, lactate and hydrogen ions accumulate, resulting in a transient metabolic acidosis [16]. Lactate is increasingly recognized not only as a marker of high metabolic stress but also as an immunometabolite that modulates immune cell differentiation, activation, migration, and function through mechanisms including receptor-mediated signaling and epigenetic regulation such as histone lactylation [23,24]. Depending on the context, lactate has been shown to exert pro- and anti-inflammatory effects [23]. Concurrently, exercise above the anaerobic threshold is accompanied by a marked increase in circulating catecholamines [53], which promote β2-adrenergic-mediated immune cell trafficking and redistribution [12,54]. Consequently, the anaerobic threshold represents a physiologically meaningful benchmark for studies investigating exercise-induced immune responses, as it coincides with marked alterations in key metabolic and neuroendocrine pathways known to influence immune function. A lactate concentration of 4 mmol/L was used to define the anaerobic threshold. With a maximum lactate concentration of 5.76 ± 1.87 mmol/L the anaerobic threshold was exceeded by the STST, and it can be concluded that the 1-minute STST is suitable as an anaerobic exercise test in non-athletes. However, although an increase in lactate concentration was observed in all participants, 4 participants did not reach a lactate concentration of 4 mmol/L, suggesting that the relative test intensity may have been lower for these individuals, resulting in a smaller contribution of anaerobic metabolism. This may be attributable to a lower pacing, as maximum heart rate and number of sit-to-stand cycles were also lower in these participants than the study cohort’s average.

The discrepancy between the intensity classification based on percentage of HRmax (moderate) and classification based on lactate concentration (anaerobic) may be attributed to several factors. The inter-individual differences in exercise intensity and physiological response, along with a possible underestimation of the peak heart rate values due to measurement limitations may have contributed to the lower HRmax-based intensity classification. Additionally, the responses are depending on the individual participants‘ physical fitness level. Furthermore, HRmax and lactate concentration reflect different aspects of exercise response, which may lead to divergent intensity categorizations, especially during short-term, high-intensity exercise. The observed tendency of decreasing glucose concentrations reflects increased utilization of glucose for energy production during exercise. Blood glucose is metabolized to generate energy, resulting in the production of lactate under anaerobic conditions [15]. The sharp increase in the lactate concentration observed 5 minutes post-exercise reflects this metabolic response. The delayed appearance of the lactate peak after the exercise can be attributed to compartmental shifts, as lactate is initially produced in the muscle cells and must be transported into the bloodstream mainly via monocarboxylate transporters (MCTs) [18].

Furthermore, it should be examined whether the exercise-induced immunological stress response is triggered by the short-term exercise. First, the decrease in the leukocyte count in the 30-minute pre-exercise phase showed that the resting phase is necessary, as getting to the study location, for example, can lead to a minor stress reaction that affects the blood cell count and can therefore lead to an overestimation of the initial values. After the test, exercise-induced leukocytosis was observed as an immediate reaction, followed by a gradual decline in leukocyte count post-exercise. Similarly, increases in lymphocytes and granulocytes immediately after the STST and their decrease afterwards could be measured. These findings indicate a redistribution of immune cells, potentially resulting from an activation of the immune system and alterations in the vascular barrier, induced by the exercise test and associated hormone release. As described previously, catecholamine and glucocorticoid release contribute to immune cell trafficking between blood and peripheral pools [3,12]. The release of catecholamines contributes to mobilization of leukocytes into the circulation by modulating expression of adhesion molecules, cytokine levels and leukocyte stiffness [12], whereas glucocorticoids influence key mechanisms underlying leukocyte diapedesis by regulating adhesion molecule expression, cytokine and chemokine production, and endothelial barrier integrity [14]. Additionally, exercise-induced increases in blood pressure and shear stress have been reported to facilitate demargination of leukocytes [13]. The opposing dynamics of the percentages of lymphocytes and granulocytes within the leukocytes reflect most likely the different reactions of lymphocytes and granulocytes to the release of catecholamines. While the proportion of lymphocytes increased immediately after exercise compared to before, the proportion of granulocytes decreased and in the post-exercise phase the proportion of lymphocytes decreased again, while the proportion of granulocytes increased. Comparable changes in lymphocyte and granulocyte counts and their relative proportions were reported by Schlagheck *et al.* for both endurance and strength training [22]. Furthermore, increases in leukocyte counts after different types of physical activity have already been demonstrated several times [9,20–22]. However, the exercise protocols in the respective studies were generally complex, involving prolonged exertion or exercise until exhaustion. In contrast, the findings of the present study demonstrate that even the STST as a simple anaerobic exertion of moderate intensity lasting only 1 minute is sufficient to induce significant alterations in blood immune cell counts. Studies examining the immediate immune responses to comparable brief exercise tests are rare. Jamurtas *et al*. conducted investigations of leukocyte responses to the wingate test, an established anaerobic test, and showed similar results for changes of leukocytes and proportions of lymphocytes and granulocytes [37]. However, they repeatedly carried out the 30-seconds sprint test with 4 minutes of recovery between sprints and performed measurements only at the end of the entire exercise test. A repeated sprint exercise was also conducted by Almeida-Neto *et al.,* showing as well increases in leukocyte, lymphocyte and neutrophil counts, but also carried out measurements at the end of the entire exercise (lasting more than 15 minutes in total) [38]. In another study, in addition to a repeated sprints test (10-second all-out) and continuous and intermittent VO2max tests, the wingate test was also administered as one-time sprint [55]. The results showed that all tests induced increases in leukocyte counts immediately post-exercise, without any differences between test protocols, demonstrating that even the short 30 s anaerobic exercise bout has significant impact on immune cell mobilization. Although these protocols induce immune responses comparable to those observed after the STST, they generally require cycle ergometers, sprint facilities or repeated maximal efforts and therefore remain primarily laboratory-based assessments. In contrast, the STST requires only a standard chair, minimal space and one minute to perform, making it substantially more feasible for routine clinical practice, rehabilitation settings and large-scale field studies while still eliciting a measurable immune-metabolic response. Moreover, the STST provides a brief, standardized maximal-effort task that minimizes the influence of prolonged pacing strategies while still eliciting a measurable metabolic challenge.

The extent of changes in different blood biomarkers has been reported to depend on exercise intensity and the participants’ training status [56,57]. Since ΔHR reflects the degree to which heart rate remains elevated 5 minutes after exercise, it may serve as an indirect indicator of exercise intensity and/or training status, whereby a greater ΔHR may indicate either a higher intensity of the test or a lower training status. In the present study, correlation analyses showed that greater ΔHR values were associated with greater ΔLactate, ΔWBC, ΔLYM, and ΔGRA. These findings support the hypothesis that both intensity and training status influence the magnitude of changes in biomarkers following exercise. However, heart rate recovery at 5 minutes post-exercise, which is commonly considered a marker of training status, did not correlate with the metabolic, immunological, or inflammatory markers at that time point, suggesting that the training status may be less determinative than exercise intensity in the observed responses.

The significant positive correlations between ΔLactate and the exercise-induced changes in leukocyte, lymphocyte and granulocyte counts indicate that the magnitude of the metabolic response to the STST is closely linked to the extent of immune cell redistribution. This is consistent with the concept that lactate is not merely a metabolic by-product but an active signaling molecule involved in the regulation of immune cell mobilization and function [18,23]. During the brief, near-maximal effort of the STST, the pronounced reliance on anaerobic glycolysis raises blood lactate above the anaerobic threshold [19], reflecting a high metabolic and sympathetic stress load; the accompanying catecholamine release is a principal driver of the rapid, demargination-mediated leukocytosis observed immediately after exercise [3,10,11]. The parallel correlations of both ΔLactate and ΔHR with the same immune parameters (Figure 4 and 5) further support the interpretation that metabolic and cardiorespiratory strain track together with acute immune cell shifts. These associations are, however, correlational and do not establish causality: lactate accumulation and leukocyte redistribution may be co-regulated by shared upstream mechanisms, notably catecholamine release and exercise-induced shear stress [11,13], rather than lactate directly recruiting immune cells into the circulation. The comparatively strong association for lymphocytes is in line with their high β-adrenergic sensitivity and preferential early mobilization [8,10]. Taken together, these findings position blood lactate as a simple, accessible readout of the internal exercise stimulus that scales with the acute immune-metabolic response to the STST, and they motivate future mechanistic studies incorporating catecholamines, cortisol and cytokines to disentangle the metabolic and endocrine contributions to exercise-induced immune regulation.

It is known that regular moderate physical activity has an anti-inflammatory effect [7]. At the same time, a single exercise bout initially leads to an acute-phase reaction and upregulation of proinflammatory cytokines [58,59]. The NLR, calculated as the simple ratio of neutrophil to lymphocyte counts, is widely used as a prognostic biomarker in various pathological conditions, including sepsis, cancer, COVID-19, and cardiovascular diseases [60]. The SII and SIRI are also considered simple inflammatory markers and are of prognostic significance for a variety of diseases [61–63]. NLR, SII and SIRI are inexpensive indices derived from the differential blood count, incorporate multiple immune cell populations and are increasingly used as markers of systemic inflammation and prognosis across a wide range of clinical settings [64]. In addition, these markers have been proposed as useful in exercise settings to characterize induced inflammatory responses [65]. Combining these readily available inflammatory indices with a simple field-based exercise protocol may facilitate broader implementation of exercise-induced immune monitoring in clinical practice and longitudinal follow-up studies. High NLR, SII and SIRI are associated with an increased state of inflammation and disturbance of the immune system. Both GLR and SII decreased significantly immediately after exercise, potentially reflecting the catecholamine-mediated redistribution of circulating leukocyte subsets. As lymphocyte counts increased proportionally more than granulocyte counts, both indices transiently declined immediately post-exercise, consistent with the exercise-induced mobilization of immune cells involved in enhanced immune surveillance [54]. These distinct dynamics, which become particularly evident when considering the changes in the relative proportions of leukocyte subsets, are summarized in the composite indices GLR and SII. A possible underlying mechanism is the greater β2-adrenergic responsiveness of circulating lymphocyte subsets, resulting in a more pronounced catecholamine-induced demargination compared to granulocytes [12,66]. Additionally, the comparatively smaller increase of granulocytes in the circulation is most plausibly explained by their lower adrenergic responsiveness relative to NK cells, cytotoxic and γδ T cells, and non-classical monocytes [67–69], compounded by a denominator effect arising from their high resting count. A second, non-exclusive factor may be their distinct margination and trafficking behavior: neutrophils are the first leukocyte subset recruited from the circulation into tissue during infection and tissue injury [70], and the size and dynamics of their marginated pool may therefore modulate the net circulating response [71]. This interpretation applies to the acute, catecholamine-driven phase [72,73]; a pronounced neutrophilia typically emerges in the hours after exercise, attributed to cortisol-mediated bone marrow release and demargination [3,10]. Following the immediate post-exercise decrease, GLR, SII and SIRI subsequently increased above baseline levels at 30 and 45 minutes post-exercise, likely reflecting the transient glucocorticoid-mediated mobilization and redistribution of immune cell subsets during recovery following the acute exercise stress stimulus. In the study by Schlagheck *et al.*, a significant increase in NLR and SII was observed too after exercise (one hour after the activity) and NLR also decreased immediately post-exercise [22]. However, the changes were observed only in the group of participants with endurance training (45-minute bicycle ergometer test at 60 % of maximal power), while the markers remained unchanged after strength training (45-minute machine training with 5 x 4 sets of 8-10 repetitions at 70 % of 1RM). A decrease in SII immediately after the activity was not observed in either group. Joisten *et al.* observed a significant increase in SII immediately up to 4 hours after physical activity (300 countermovement jumps with maximal effort in 40 minutes), but also no decrease in SII immediately after exercise [36]. The observations immediately after exercise, which are inconsistent with the literature, may be related to differences in the type of exercise (duration and intensity), as these affect the acute-phase response and changes in blood cells. More intense activities are associated with a greater increase in cytokines such as IL-6 [2] and with higher cell counts, particularly of lymphocytes [8]. In contrast, exercise-induced neutrophilia depends more on the duration than the intensity of the activity [8].

The present findings suggest that the STST complements existing functional field tests by providing a standardized exercise challenge capable of inducing measurable metabolic and immune responses. It combines the practicality and accessibility of a clinical field test with the ability to induce measurable metabolic and immunological responses, thereby expanding its potential applications beyond functional performance assessment. Furthermore, it demonstrates translational potential for clinical settings as the results may not only provide a norm for the physiological response to exercise, but also comparative data for evaluating the immune-metabolic response to the STST in clinical populations, where the test may provide utility for monitoring immune function in chronic diseases or recovery after illness. Potential target populations include individuals with metabolic-associated fatty liver disease (MAFLD), inflammatory bowel disease (IBD), post-acute infection syndromes, cancer-related fatigue or other chronic conditions in whom maximal laboratory exercise testing is often impractical or not feasible. Furthermore, the simplicity of the protocol makes repeated assessments in primary care, outpatient rehabilitation and telemedicine settings conceivable. Future studies are warranted to determine whether this protocol could help identify altered immune responses to exercise and differentiate physiological from suppressed immune responses. The potential clinical and non-clinical applications of the STST as a tool based on the aforementioned previous evaluations and the findings of this study are summarized in Table 3.

**Table 3.** Potential applications and future directions of the 1-minute sit-to-stand test (STST) as a tool in clinical and non-clinical settings. Each application indicates the current supporting evidence and, where applicable, is anchored to references already cited in the manuscript.

| Potential advantage of the 1-min STST | Clinical applications | Non-clinical (research) applications |
| --- | --- | --- |
| **Simple, standardized equipment-free field test** | Validated, low-cost measure of functional exercise capacity that requires no laboratory infrastructure and is feasible at the bedside or in outpatient settings (established) [26,27,29]; population-based and disease-specific reference values are available to aid interpretation [44-47]. | Provides a standardized, reproducible exercise stimulus for exercise-immunology and immunometabolism research in field or resource-limited settings (proposed) [24,65]. |
| **Elicits measurable, reproducible metabolic and immune responses** | Demonstrates significant post-exercise increases in lactate, leukocyte, lymphocyte and granulocyte counts and modulation of GLR, SII and SIRI (this study), consistent with the established acute exercise leukocytosis [3,8,10,20]. This may support follow-up evaluation in populations such as post-acute infection syndromes without inducing symptom exacerbation (proposed). | Enables mechanistic study of lactate-immune interactions and acute immune-metabolic coupling (this study) [18,23,52]. |
| **Potential surrogate for / complement to laboratory-based exercise protocols** | Correlates with the 6-min walk test in COPD and in healthy adults and avoids the learning effects and infrastructure of walk- or step-based tests (established) [27,29-34]; direct validation as an immune-metabolic surrogate is still required (proposed). | Offers a practical alternative for exercise-immunology field studies when treadmill or cycle ergometry is unavailable or impractical (proposed) [24]. |
| **Advantages over other established simple field tests (6-min walk test, step test)** | Feasible even in individuals with limited physical fitness, unlike the Chester step test, which is physically demanding and cannot be completed by some individuals; also needs only a small space and less time than the 6-min walk test, which requires a ~30-m hallway and a familiarization trial; usable across a wider range of in- and out-of-hospital settings, including primary care and tele-rehabilitation (established) [26-28]. Provides estimates of functional exercise capacity comparable to the 6-min walk test despite a different cardiorespiratory demand (established) [27,29,30]. | Offers a more standardized stimulus than the self-paced 6-min walk test, which shows a documented learning effect [31,32], or the Chester step test, an incremental test whose exercise duration and achieved workload depend on the participant's performance and termination point, so that participants are exposed to different cumulative stimuli despite a standardized protocol [33,34]; by contrast, the STST's brief, fixed-duration maximal-effort format minimizes pacing effects and provides a more comparable acute immune-metabolic stimulus (this study / proposed) [26-30]. |
| **Broad applicability across populations** | Feasible in patients with reduced physical function, including COPD, long/post-COVID and older adults (established) [27-29,74,75]; extension to MAFLD, cancer-related fatigue and post-acute infection syndromes (proposed). | Allows inclusion of heterogeneous cohorts with varying fitness levels in immunology and physiology studies (proposed). |
| **Low burden enables repeated and longitudinal measurement** | Suitable for monitoring functional change over time in cardiopulmonary rehabilitation (established) [26]; longitudinal monitoring of immune-metabolic recovery after illness (proposed). | Supports repeated assessment of immune-metabolic adaptations in intervention and longitudinal research designs (proposed) [42]. |

**Supporting evidence:** (established) = supported by prior peer-reviewed literature on the STST or on acute exercise immunology (cited); (this study) = demonstrated by the present findings; (proposed) = hypothesis or future direction not yet directly tested for the STST. Bracketed numbers refer to the manuscript reference list. COPD: chronic obstructive pulmonary disease, GLR: granulocyte-to-lymphocyte ratio, MAFLD: metabolic-associated fatty liver disease, SII: systemic immune-inflammation index, SIRI: systemic inflammatory response index, STST: sit-to-stand test, 6-min walk test: six-minute walk test.

Besides several strengths of the study such as the combined analysis of metabolic and immunological biomarkers or the application of hemoconcentration correction, several additional limitations beyond those already mentioned, should be acknowledged. Firstly, the study population was limited to healthy, young adults, restricting the generalizability of the findings, especially to clinical populations or older adults. Secondly, the moderate sample size of 26 participants could affect generalizability as well. Thirdly, heart rate recovery data as well as height and weight were only available for 20 of 26 participants. Additionally, further standardized information about the participants’ physical activity level and information on menstrual cycle phase (for female participants) were not recorded, representing potential sources of confounding. Furthermore, GRA were used instead of neutrophil counts to calculate NLR (GLR in the present study), SII and SIRI. Although neutrophils account for the majority of circulating granulocytes, these indices should therefore be interpreted as approximations of the conventional NLR, SII and SIRI. Lastly, the inclusion of additional biomarkers, such as cytokines, catecholamines or cortisol would have provided further mechanistic insight into the observed immune-metabolic responses and strengthened the interpretation of results. However, the aim of our study was focused on the examination of the effects of the STST on immunologic and metabolic stress indices. Based on our results future mechanism-based studies should address the exerkine and hormone responses as well as interactions with immune cell mobilization.

## 5 Conclusion

Previous studies have almost exclusively applied the STST as a measure of functional exercise capacity or physical performance across different patient and non-patient populations [26,27,74–76]. The present study extends this concept by demonstrating that the STST also elicits a reproducible metabolic and immunological response represented by rapid increase in blood lactate concentration immediately after exercise, as well as leukocytosis characterized by increased lymphocyte and granulocyte counts and modulated inflammatory indices. Thus, beyond its established role as a functional assessment tool, the STST may also represent a standardized field-based model for investigating acute immune-metabolic responses under a brief, standardized maximal exercise stimulus.

## Data availability

Data as well as Pythons scripts are available in the Open Science Framework (OSF) repository: https://doi.org/10.17605/OSF.IO/BNPEV.

## Author contributions

**Wiebke Röhr:** Conceptualization, Methodology, Formal analysis, Investigation, Data curation, Visualization, Writing – Original Draft. **Rebecca Simon:** Methodology, Writing – Review & Editing. **Sebastian Kirschke:** Formal analysis, Visualization. **Isabell von Loga:** Writing – Review & Editing. **David Putrino:** Writing – Review & Editing. **Wilhelm Bloch:** Writing – Review & Editing. **Philipp A. Reuken:** Writing – Review & Editing. **Diana Dudziak:** Writing – Review & Editing. **Anna P. Kipp:** Conceptualization, Supervision, Writing – Review & Editing. **Andreas Stallmach:** Writing – Review & Editing. **Christian Puta:** Conceptualization, Supervision, Writing – Review & Editing, Funding acquisition.

All authors approved the final version of the manuscript submitted for publication.

## References

1 Phelps, C. M. and Meisel, M. The immunology of exercise: Mechanisms, mediators, and therapeutic opportunities. Immunity (2026). 10.1016/j.immuni.2026.04.016

2 Moldoveanu, A. I., Shephard, R. J. and Shek, P. N. The cytokine response to physical activity and training. Sports Med. 31, 115–144 (2001). 10.2165/00007256-200131020-00004

3 Peake, J. M., Neubauer, O., Walsh, N. P. and Simpson, R. J. Recovery of the immune system after exercise. J Appl Physiol (1985) 122, 1077–1087 (2017). 10.1152/japplphysiol.00622.2016

4 Pedersen, B. K. Exercise and cytokines. Immunol. Cell Biol. 78, 532–535 (2000). 10.1111/j.1440-1711.2000.t01-11-.x

5 Petersen, A. M. and Pedersen, B. K. The anti-inflammatory effect of exercise. J Appl Physiol (1985) 98, 1154–1162 (2005). 10.1152/japplphysiol.00164.2004

6 Aronson, D. et al. C-Reactive protein is inversely related to physical fitness in middle-aged subjects. Atherosclerosis 176, 173–179 (2004). 10.1016/j.atherosclerosis.2004.04.025

7 Pischon, T., Hankinson, S. E., Hotamisligil, G. S., Rifai, N. and Rimm, E. B. Leisure-time physical activity and reduced plasma levels of obesity-related inflammatory markers. Obes Res 11, 1055–1064 (2003). 10.1038/oby.2003.145

8 Gabriel, H. and Kindermann, W. The Acute Immune Response to exercise: What Does It Mean? Int. J. Sports Med. 18, S28–S45 (1997). 10.1055/s-2007-972698

9 McCarthy, D. A. et al. Studies on the immediate and delayed leukocytosis elicited by brief (30-min) strenuous exercise. Eur. J. Appl. Physiol. Occup. Physiol. 64(6), 513–517 (1992). 10.1007/BF00843760

10 McCarthy, D. A. and Dale, M. M. The leucocytosis of exercise. A review and model. Sports Med 6, 333–363 (1988). 10.2165/00007256-198806060-00002

11 Zouhal, H., Jacob, C., Delamarche, P. and Gratas-Delamarche, A. Catecholamines and the effects of exercise, training and gender. Sports Med. 38, 401–423 (2008). 10.2165/00007256-200838050-00004

12 Ince, L. M., Weber, J. and Scheiermann, C. Control of Leukocyte Trafficking by Stress-Associated Hormones. Front Immunol 9, 3143 (2018). 10.3389/fimmu.2018.03143

13 Shephard, R. J. Adhesion molecules, catecholamines and leucocyte redistribution during and following exercise. Sports Med 33, 261–284 (2003). 10.2165/00007256-200333040-00002

14 Zielinska, K. A., Van Moortel, L., Opdenakker, G., De Bosscher, K. and Van den Steen, P. E. Endothelial Response to Glucocorticoids in Inflammatory Diseases. Front Immunol 7, 592 (2016). 10.3389/fimmu.2016.00592

15 Hargreaves, M. and Spriet, L. L. Skeletal muscle energy metabolism during exercise. Nat Metab 2, 817–828 (2020). 10.1038/s42255-020-0251-4

16 Sahlin, K., Tonkonogi, M. and Soderlund, K. Energy supply and muscle fatigue in humans. Acta Physiol Scand 162, 261–266 (1998). 10.1046/j.1365-201X.1998.0298f.x

17 Spriet, L. L., Howlett, R. A. and Heigenhauser, G. J. An enzymatic approach to lactate production in human skeletal muscle during exercise. Med. Sci. Sports Exerc. 32, 756–763 (2000). 10.1097/00005768-200004000-00007

18 Li, X. et al. Lactate metabolism in human health and disease. Signal Transduct Target Ther 7, 305 (2022). 10.1038/s41392-022-01151-3

19 Wasserman, K., Whipp, B. P., Koyl, S. N. and Beaver, W. L. Anaerobic threshold and respiratory gas exchange during exercise. J. Appl. Physiol. 35(2), 236–243 (1973). 10.1152/jappl.1973.35.2.236

20 Hansen, J. B., Wilsgård, L. and Osterud, B. Biphasic changes in leukocytes induced by strenuous exercise. Eur. J. Appl. Physiol. Occup. Physiol. 62, 157–161 (1991). 10.1007/bf00643735

21 Gabriel, H., Schwarz, L., Born, P. and Kindermann, W. Differential mobilization of leucocyte and lymphocyte subpopulations into the circulation during endurance exercise. Eur. J. Appl. Physiol. Occup. Physiol. 65, 529–534 (1992). 10.1007/bf00602360

22 Schlagheck, M. L. et al. Cellular immune response to acute exercise: Comparison of endurance and resistance exercise. Eur J Haematol 105, 75–84 (2020). 10.1111/ejh.13412

23 Llibre, A., Kucuk, S., Gope, A., Certo, M. and Mauro, C. Lactate: A key regulator of the immune response. Immunity 58, 535–554 (2025). 10.1016/j.immuni.2025.02.008

24 Nieman, D. C. and Pence, B. D. Exercise immunology: Future directions. J Sport Health Sci 9, 432–445 (2020). 10.1016/j.jshs.2019.12.003

25 Faude, O., Kindermann, W. and Meyer, T. Lactate threshold concepts: how valid are they? Sports Med. 39, 469–490 (2009). 10.2165/00007256-200939060-00003

26 Bohannon, R. W. and Crouch, R. 1-Minute Sit-to-Stand Test: SYSTEMATIC REVIEW OF PROCEDURES, PERFORMANCE, AND CLINIMETRIC PROPERTIES. J Cardiopulm Rehabil Prev 39, 2–8 (2019). 10.1097/HCR.0000000000000336

27 Reychler, G. et al. One minute sit-to-stand test is an alternative to 6MWT to measure functional exercise performance in COPD patients. Clin Respir J 12, 1247–1256 (2018). 10.1111/crj.12658

28 Sevillano-Castaño, A. I. et al. Test–Retest Reliability and Minimal Detectable Change in Chester Step Test and 1-Minute Sit-to-Stand Test in Long COVID Patients. Applied Sciences 13 (2023). 10.3390/app13148464

29 Ozalevli, S., Ozden, A., Itil, O. and Akkoclu, A. Comparison of the Sit-to-Stand Test with 6 min walk test in patients with chronic obstructive pulmonary disease. Respir Med 101, 286–293 (2007). 10.1016/j.rmed.2006.05.007

30 Gurses, H. N., Zeren, M., Denizoglu Kulli, H. and Durgut, E. The relationship of sit-to-stand tests with 6-minute walk test in healthy young adults. Medicine (Baltimore) 97, e9489 (2018). 10.1097/MD.0000000000009489

31 Wu, G., Sanderson, B. and Bittner, V. The 6-minute walk test: how important is the learning effect? Am Heart J 146, 129–133 (2003). 10.1016/s0002-8703(03)00119-4

32. ATS Committee on Proficiency Standards for Clinical Pulmonary Function Laboratories. ATS statement: guidelines for the six-minute walk test. Am J Respir Crit Care Med 166, 111–117 (2002). 10.1164/ajrccm.166.1.at1102

33 Buckley, J. P., Sim, J., Eston, R. G., Hession, R. and Fox, R. Reliability and validity of measures taken during the Chester step test to predict aerobic power and to prescribe aerobic exercise. Br J Sports Med 38, 197–205 (2004). 10.1136/bjsm.2003.005389

34 Sykes, K. and Roberts, A. The Chester step test—a simple yet effective tool for the prediction of aerobic capacity. Physiotherapy 90, 183–188 (2004). 10.1016/j.physio.2004.03.008

35 McKay, A. K. A. et al. Defining Training and Performance Caliber: A Participant Classification Framework. Int J Sports Physiol Perform 17, 317–331 (2022). 10.1123/ijspp.2021-0451

36 Joisten, N. et al. Aqua cycling for immunological recovery after intensive, eccentric exercise. Eur J Appl Physiol 119, 1369–1375 (2019). 10.1007/s00421-019-04127-4

37 Jamurtas, A. Z. et al. The Effects of Acute Low-Volume HIIT and Aerobic Exercise on Leukocyte Count and Redox Status. J Sports Sci Med 17, 501–508 (2018).

38 Almeida-Neto, P. F. d., et al. Leukocyte response to repeated sprint exercise: Differences between adolescent and adult athletes. Sports Medicine and Health Science (2025). 10.1016/j.smhs.2025.05.003

39 Gaubitz, M. et al. Efficacy and safety of nicoboxil/nonivamide ointment for the treatment of acute pain in the low back - A randomized, controlled trial. Eur J Pain 20, 263–273 (2016). 10.1002/ejp.719

40 Dill, D. B. and Costill, D. L. Calculation of percentage changes in volumes of blood, plasma, and red cells in dehydration. J. Appl. Physiol. 37(2), 247–248 (1974). 10.1152/jappl.1974.37.2.247

41 Matomäki, P., Kainulainen, H. and Kyröläinen, H. Corrected whole blood biomarkers - the equation of Dill and Costill revisited. Physiol. Rep. 6, e13749 (2018). 10.14814/phy2.13749

42 Steidten, T. et al. Impact of different concurrent training sequencing schemes on overnight systemic immunological regulation in adolescent athletes. Front. Physiol. 16, 1392946 (2025). 10.3389/fphys.2025.1392946

43 Nes, B. M., Janszky, I., Wisløff, U., Støylen, A. and Karlsen, T. Age-predicted maximal heart rate in healthy subjects: The HUNT fitness study. Scand. J. Med. Sci. Sports 23, 697–704 (2013). 10.1111/j.1600-0838.2012.01445.x

44 Vilarinho, R., Montes, A. M., Noites, A., Silva, F. and Melo, C. Reference values for the 1-minute sit-to-stand and 5 times sit-to-stand tests to assess functional capacity: a cross-sectional study. Physiotherapy 124, 85–92 (2024). 10.1016/j.physio.2024.01.004

45 Strassmann, A. et al. Population-based reference values for the 1-min sit-to-stand test. Int J Public Health 58, 949–953 (2013). 10.1007/s00038-013-0504-z

46 Otto-Yanez, M. et al. One-minute sit-to-stand test: Reference values for the Chilean population. PLoS One 20, e0317594 (2025). 10.1371/journal.pone.0317594

47 Furlanetto, K. C. et al. Reference Values for 7 Different Protocols of Simple Functional Tests: A Multicenter Study. Arch Phys Med Rehabil 103, 20–28.e25 (2022). 10.1016/j.apmr.2021.08.009

48 Garber, C. E. et al. American College of Sports Medicine position stand. Quantity and quality of exercise for developing and maintaining cardiorespiratory, musculoskeletal, and neuromotor fitness in apparently healthy adults: guidance for prescribing exercise. Med Sci Sports Exerc 43, 1334–1359 (2011). 10.1249/MSS.0b013e318213fefb

49 Daanen, H. A., Lamberts, R. P., Kallen, V. L., Jin, A. and Van Meeteren, N. L. A systematic review on heart-rate recovery to monitor changes in training status in athletes. Int. J. Sports Physiol. Perform. 7, 251–260 (2012). 10.1123/ijspp.7.3.251

50 Römer, C. and Wolfarth, B. Heart Rate Recovery (HRR) Is Not a Singular Predictor for Physical Fitness. Int. J. Env. Res. Public Health 20 (2022). 10.3390/ijerph20010792

51 Sales, M. M. et al. An integrative perspective of the anaerobic threshold. Physiol Behav 205, 29–32 (2019). 10.1016/j.physbeh.2017.12.015

52 O’Neill, L. A., Kishton, R. J. and Rathmell, J. A guide to immunometabolism for immunologists. Nat Rev Immunol 16, 553–565 (2016). 10.1038/nri.2016.70

53 Urhausen, A., Weiler, B., Coen, B. and Kindermann, W. Plasma catecholamines during endurance exercise of different intensities as related to the individual anaerobic threshold. Eur J Appl Physiol Occup Physiol 69, 16–20 (1994). 10.1007/bf00867921

54 Simpson, R. J. et al. Exercise and adrenergic regulation of immunity. Brain Behav Immun 97, 303–318 (2021). 10.1016/j.bbi.2021.07.010

55 Minuzzi, L. G. et al. Acute Hematological and Inflammatory Responses to High-intensity Exercise Tests: Impact of Duration and Mode of Exercise. Int J Sports Med 38, 551–559 (2017). 10.1055/s-0042-117723

56 Neves, P. et al. Acute effects of high- and low-intensity exercise bouts on leukocyte counts. J Exerc Sci Fit 13, 24–28 (2015). 10.1016/j.jesf.2014.11.003

57 Fortunato, A. K. et al. Strength Training Session Induces Important Changes on Physiological, Immunological, and Inflammatory Biomarkers. J. Immunol. Res. 2018, 9675216 (2018). 10.1155/2018/9675216

58 Ostrowski, K. et al. A trauma-like elevation of plasma cytokines in humans in response to treadmill running. J. Physiol. 513 ( Pt 3), 889–894 (1998). 10.1111/j.1469-7793.1998.889ba.x

59 Ostrowski, K., Rohde, T., Asp, S., Schjerling, P. and Pedersen, B. K. Pro- and anti-inflammatory cytokine balance in strenuous exercise in humans. J. Physiol. 515 ( Pt 1), 287– 291 (1999). 10.1111/j.1469-7793.1999.287ad.x

60 Buonacera, A., Stancanelli, B., Colaci, M. and Malatino, L. Neutrophil to Lymphocyte Ratio: An Emerging Marker of the Relationships between the Immune System and Diseases. Int J Mol Sci 23 (2022). 10.3390/ijms23073636

61 Fois, A. G. et al. The Systemic Inflammation Index on Admission Predicts In-Hospital Mortality in COVID-19 Patients. Molecules 25 (2020). 10.3390/molecules25235725

62 Liu, X. et al. The Combination of Hemogram Indexes to Predict Exacerbation in Stable Chronic Obstructive Pulmonary Disease. Front Med (Lausanne) 7, 572435 (2020). 10.3389/fmed.2020.572435

63 Hu, B. et al. Systemic immune-inflammation index predicts prognosis of patients after curative resection for hepatocellular carcinoma. Clin Cancer Res 20, 6212–6222 (2014). 10.1158/1078-0432.CCR-14-0442

64 Islam, M. M., Satici, M. O. and Eroglu, S. E. Unraveling the clinical significance and prognostic value of the neutrophil-to-lymphocyte ratio, platelet-to-lymphocyte ratio, systemic immune-inflammation index, systemic inflammation response index, and delta neutrophil index: An extensive literature review. Turk J Emerg Med 24, 8–19 (2024). 10.4103/tjem.tjem_198_23

65 Walzik, D., Joisten, N., Zacher, J. and Zimmer, P. Transferring clinically established immune inflammation markers into exercise physiology: focus on neutrophil-to-lymphocyte ratio, platelet-to-lymphocyte ratio and systemic immune-inflammation index. Eur J Appl Physiol 121, 1803–1814 (2021). 10.1007/s00421-021-04668-7

66 Pedersen, B. K. and Hoffman-Goetz, L. Exercise and the immune system: regulation, integration, and adaptation. Physiol Rev 80, 1055–1081 (2000). 10.1152/physrev.2000.80.3.1055

67 Landmann, R. Beta-adrenergic receptors in human leukocyte subpopulations. Eur J Clin Invest 22 Suppl 1, 30–36 (1992).

68 Dimitrov, S., Lange, T. and Born, J. Selective mobilization of cytotoxic leukocytes by epinephrine. J Immunol 184, 503–511 (2010). 10.4049/jimmunol.0902189

69 Graff, R. M. et al. β(2)-Adrenergic receptor signaling mediates the preferential mobilization of differentiated subsets of CD8+ T-cells, NK-cells and non-classical monocytes in response to acute exercise in humans. Brain Behav Immun 74, 143–153 (2018). 10.1016/j.bbi.2018.08.017

70 Kolaczkowska, E. and Kubes, P. Neutrophil recruitment and function in health and inflammation. Nat Rev Immunol 13, 159–175 (2013). 10.1038/nri3399

71 Summers, C. et al. Neutrophil kinetics in health and disease. Trends Immunol 31, 318–324 (2010). 10.1016/j.it.2010.05.006

72 Simpson, R. J., Kunz, H., Agha, N. and Graff, R. Exercise and the Regulation of Immune Functions. Prog Mol Biol Transl Sci 135, 355–380 (2015). 10.1016/bs.pmbts.2015.08.001

73 Walsh, N. P. et al. Position statement. Part one: Immune function and exercise. Exerc Immunol Rev 17, 6–63 (2011).

74 Cruz-Montecinos, C. et al. Which Sit-to-Stand Test Best Differentiates Functional Capacity in Older People? Am J Phys Med Rehabil 103, 925–928 (2024). 10.1097/PHM.0000000000002504

75 Nunez-Cortes, R. et al. Use of sit-to-stand test to assess the physical capacity and exertional desaturation in patients post COVID-19. Chron Respir Dis 18, 1479973121999205 (2021). 10.1177/1479973121999205

76 Roldan-Jimenez, C., Bennett, P. and Cuesta-Vargas, A. I. Muscular Activity and Fatigue in Lower-Limb and Trunk Muscles during Different Sit-To-Stand Tests. PLoS One 10, e0141675 (2015). 10.1371/journal.pone.0141675

